# ‘Doing nothing’ is simply not an option: why framing of choices matters in surgical shared decision-making

**DOI:** 10.1101/2022.07.27.22278115

**Authors:** Agata Ludwiczak, Timothy Stephens, John Prowle, Rupert Pearse, Magda Osman

## Abstract

**Background:** In the context of high-risk surgery, shared decision-making (SDM) can be hindered by misalignment in expectations regarding the likely surgical outcomes between patients and clinicians. This study investigates the extent of this misalignment in high-risk patients and doctors involved in perioperative care, its’ impact on treatment choices, and its’ amenability to interventions that encourage perspective taking.

**Methods:** High-risk patients (N = 55) and doctors involved in perioperative care (N = 54) were asked to consider one of three clinical scenarios: ischaemic heart disease, colorectal cancer, or osteoarthritis of the left hip. They reported on their expectations regarding short- and long-term outcomes of different treatment options available in these scenarios. Participants were initially asked to consider the scenarios from their own perspective as a patient/clinician, and then to adopt the perspective of the other side. After stating their expectations, participants were required to choose between surgical or non-surgical treatment.

**Results:** Systematic misalignment in expectations between high-risk patients and doctors was observed, with patients expecting better surgical outcomes compared to clinicians. Despite this misalignment, in both groups surgical treatment was strongly preferred. Willingness to consider the non-surgical option was only observed when this option offered a chance to change the undesirable ‘status quo’.

**Conclusion:** When high-risk surgery is discussed, a non-surgical option may be viewed as ‘doing nothing’, reducing the sense of agency and control. This biases the decision-making process, regardless of the expectations doctors and patients might have about the outcomes of surgery. Thus, to improve SDM and to increase patients’ agency and control over decisions about their care, we advocate framing the non-surgical treatment options in a way that emphasises action, agency, and change.

**Highlights:**

- Misalignment in expectations regarding treatment outcomes between high-risk surgical patients and their clinicians has been identified in this study, with patients expecting more positive outcomes from surgery than doctors
- Despite misalignment, treatment choices were similar for patients and clinicians
- Framing the treatment choice as ‘doing something’ (i.e. surgery) vs. ‘doing nothing’ seemed to drive the preference for surgery in both groups
- To increase patients’ agency and control over decisions about surgery, the framing of their options should be targeted for improvement

## Introduction

The value of the shared decision-making (SDM) approach is in helping patients and doctors make informed decisions about treatment, while taking into account the goals and priorities of a patient^1^. This is particularly important in the context of high-risk surgery, where serious medical complications leading to long-term decline in health and quality of life are relatively common^2-4^. Despite the benefits of this approach, SDM is still relatively underused in the context of high-risk surgery^5^. One of the barriers to achieving successful SDM is the lack of understanding of the expectations doctors and patients have about the outcomes of surgery, and how these expectations are aligned/misaligned. The aim of this study was to address this barrier by identifying the areas of alignment/misalignment, and by exploring the potential of perspective taking to bring about mutual understanding between patients and doctors.

For some time now, SDM has been advocated as an ideal model of treatment decision-making in patient-doctor consultations, and a means of improving the way in which patients are assisted in making informed decisions^6^. Conceptualised as an alternative to ‘paternalistic’ decision-making model in medicine (where the doctor has the ultimate responsibility for the decision that is made), SDM aims to incorporate patients’ values and goals into the consultation process, resulting in mutual agreement regarding the best treatment option^7^. While this approach has been shown to benefit both patients and doctors via improved information sharing and increased knowledge^8, 9^,there is still a lack of clear guidance about how to accomplish SDM in routine surgical consultations^10^. Many different shared decision-making tools are available, but the evidence regarding their efficacy (as measured by patient satisfaction scores) is mixed^11, 12^. One reason that has been offered for this is that there is no established alignment in the patient and doctor’s views to begin with, in order for a shared decision-making process to proceed^12-14^.

One possible solution to this, which has yet to be examined in a systematic empirical way, is to use a perspective taking approach^15-17^. In order to enhance effective communication in general, encouraging those involved in a dialogue (within a dyad) to assume the perspective of the other can help to expose some of the differences as well as the shared views, beliefs, and preferences that the dyad have^18^. By extension, in patient-clinician communication, some have suggested that a perspective-taking approach can help doctors increase their understanding of the needs of patients, as well as enhance empathic concern for the experiences that they have^16, 17^. However, thus far, the empirical work examining the use of perspective taking in patient-doctor consultations is extremely limited^19^.

Michie, Miles, and Weinman^19^ reviewed studies of patients with a chronic illness consulting health professionals. The review generated a total of 30 eligible empirical studies examining a variety of approaches, of which 10 studies specifically incorporated methods in which doctors took on a “patient-centred” perspective. This involved health professionals encouraging the patient to explicitly detail their perceptions on what their goals and preferences were regarding treatment. The findings of the review revealed that patient satisfaction was higher when doctors assumed a patient-centred perspective, compared with studies in which this was not adopted. In addition, this approach was associated with better physical health outcomes. If the initial findings from this and other reviews^20^ suggest that subjective (satisfaction) and objective (health) outcomes are improved by increasing the opportunity for health professionals to appreciate a patients’ perspective during the consultation process, then subjective and objective outcomes may be further enhanced by giving patients an opportunity to adopt the role of health professionals. As mentioned, the alignment of views in a shared decision-making process requires that *both* parties in the dyadic set up have a closer shared understanding of goals, preferences and risk, so that the decision a patient makes is well informed and truly reflects their views.

To build on previous work, and to help advance ways in which to improve SDM tools in high-risk patient populations, the aim of this study was to examine the expectations of high-risk patients and their doctors regarding the potential outcomes of surgery, in order to determine where they are most aligned, where they are potentially misaligned, and what impact this has on decisions that are made. We also investigated if changing perspective has an impact on alignment between patients and doctors, and whether it can be used to support better SDM processes.

To achieve the aims of this study, a novel online experiment was created, which explored the clinical decision-making process in three different disease scenarios (ischaemic heart disease, colorectal cancer, and osteoarthritis in left hip). The experiment investigated the expectations that are formed in such scenarios regarding the short- and long-term outcomes of different treatment options, and the effects changing perspective has on these expectations. To obtain the viewpoint of both patients and doctors, the questionnaire was distributed among doctors involved in the perioperative care, and a group of older lay participants (≥65 years old) with comorbidities that would class them as “high-risk” should they require surgery. The main questions this exploratory experimental set-up was designed to answer were: 1) to what extent are high-risk patients and their doctors systematically misaligned in their short- and long-term estimates of the likelihood of experiencing different outcomes that affect their quality of life following surgery (engaging in important activities, feeling pain and discomfort, feeling depressed of anxious, facing health complications), and 2) is this misalignment observed in the choices that doctors and patients face?

## Methods

### Participants

Fifty-four clinicians and fifty-five lay participants from the UK took part in this study.

All lay participants (23 females) were older than 65 years of age, and all had a Charlson Co-morbidity Index^21^ (CCI) greater than or equal to four (M=5.71; SD=2.14). Age (≥65) and the CCI score (≥4) constituted specific recruitment criteria, to obtain a sample that would have the same health characteristics as high-risk surgical patients. Online research recruitment platform *Prolific*.*co* was used for distributing the survey among the population of interest across the UK. In line with the platform’s policies, participants were paid £9 (approx. $12.33) for completing the study.

To gain the clinicians’ perspective, 26 surgeons, 25 anaesthetists, and 3 other doctors taking care of surgical patients (ICU) were recruited for this study (18 females). They were typically younger than 60 years of age (with four exceptions). Professional networks and word of mouth were used to distribute the survey among doctors involved in the care of surgical patients in London and Liverpool (UK).

As the expectations regarding treatment outcomes are likely to depend on the underlying health problem and treatment options available, participants were randomly assigned to one of three conditions, which differed with respect to the clinical scenario that was described in the online questionnaire. In the Coronary Artery Bypass Grafting (CABG) condition (22 lay participants, 22 doctors), participants were presented with a story of a patient suffering from ischaemic heart disease. In this scenario coronary artery bypass grafting was the surgical alternative to be considered, while non-surgical option involved stents or management with medications. In the Colorectal Cancer Surgery (ColRec) condition (16 lay participants, 17 doctors), participants made a choice between a surgery to remove bowel cancer vs. palliative care. In the Hip Replacement Surgery condition (Ortho) (17 lay participants, 15 doctors), osteoarthritis in the left hip was described, with hip replacement surgery vs. management with physiotherapy, walking aids and painkillers as alternatives to be considered. The exact scenarios presented in different conditions are available in the Supplementary Materials.

As the subject matter of the online survey (i.e. high-risk surgery) could be distressing for participants facing a similar choice in real life, only people not under surgical review at the time of the study were invited to take part. The study received ethical approval from the London Stanmore Research Ethics Committee (19/LO/1956). Informed consent was obtained from all participants through an online form. Participants were made aware that they could withdraw from the study at any point.

### Materials

The main component of this study was an online experiment, programmed using Qualtrics Online Survey platform^22^. The diagram of the task is presented in Figure 1. The experiment consisted of two tasks: Estimation and Perspective Taking. In both tasks participants were presented with a hypothetical scenario involving a medical condition. For a given scenario, two treatment options were presented: one which involved a surgical procedure, and one which involved a non-surgical alternative. For both options, participants were required to estimate (on a scale from 0 to 100) the likelihood of the following outcomes: *(1) ability to engage in normal activities, (2) feeling pain and discomfort, (3) feeling depressed, (4) experiencing health complications*. Each likelihood estimate was made at six timepoints: immediately after treatment, 1, 3, 6, 9 and 12 months after treatment. After considering the likely outcomes of the surgical and non-surgical alternatives, participants had to make a choice between the two treatment options.

**Figure 1.**
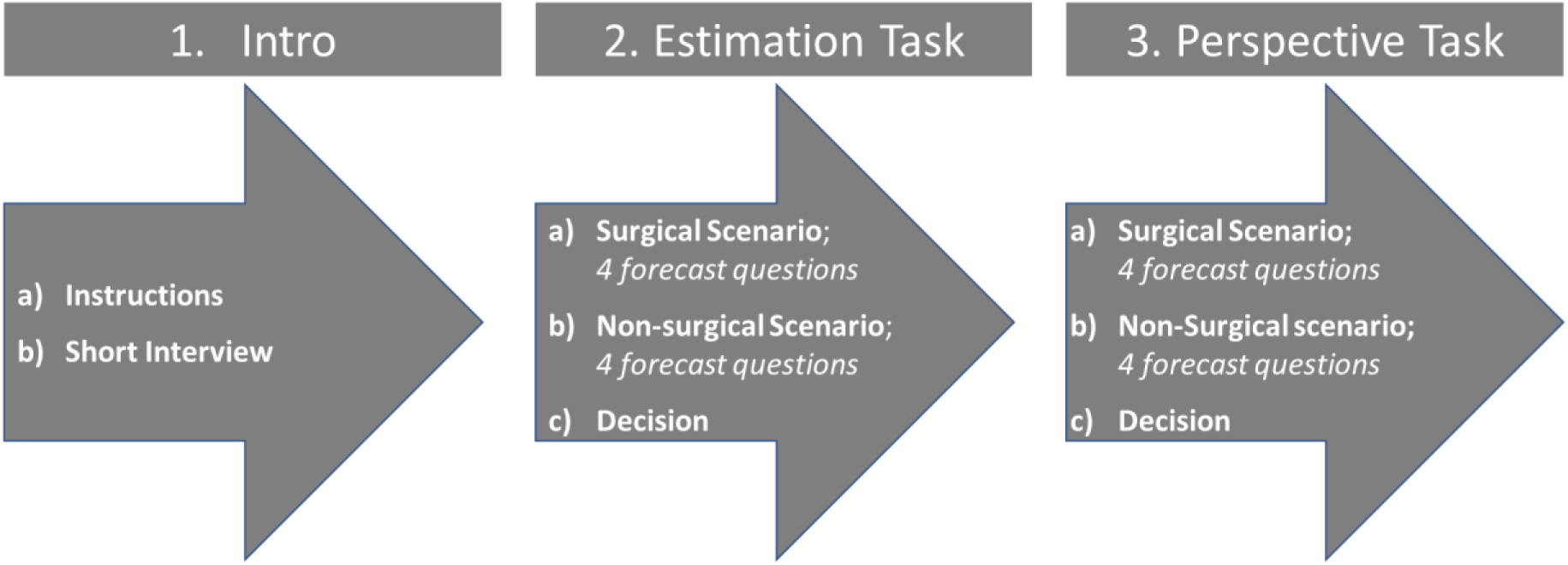
Schematic representation of the forecasting task completed by participants

#### Estimation Task

In the Estimation task lay participants were asked to consider the hypothetical disease scenario as if it was them (with their age and comorbidities) who faced the choice between the surgical and non-surgical treatment alternatives. To make this task resemble a real-life consultation, lay participants watched short videos of the experimenter delivering information about different treatment options verbally, as a doctor would. As for doctors, in the Estimation task they were asked to consider the disease scenario as if it was them who were advising a patient with this condition. Information about treatment outcomes was provided in text format for this group.

#### Perspective Taking Task

Once the Estimation task was completed, participants were informed that they would be repeating the same task, but this time they were to ‘switch roles’ and assume the role of either a ‘patient’ (if the participant was a doctor), or a ‘doctor’ (in the case of lay participants). Participants were asked to call to mind their experiences of patients/doctors and imagine themselves in the role, by considering the life they might lead, the physical and mental experiences they might have, and how they might feel. Following this prompt, participants completed the exact same procedure as in the Estimation task. For lay participants pretending to be doctors the information about treatment options was presented in text format, whereas doctors saw it delivered verbally by the experimenter, to make this experience more similar to a real-life consultation.

### Procedure

Participants accessed the online questionnaire via a link provided through the *Prolific* platform (lay participants), or through an email with an invitation to take part (doctors). They first completed the consent form, followed by a short section asking about their typical daily activities (pre-COVID-19 pandemic). This was followed by the Estimation task, and then Perspective Taking task.

#### Data Analysis

To investigate the misalignment in expectations regarding surgical and non-surgical outcomes between patients and doctors, beta regression was performed using a ‘glmmTMB’ package^23^ for statistical software R^24^. This method was selected due to its suitability for analysis of data bounded on two sides (in our case 0 and 100), with repeated measures design and unequal number of participants in each group^25, 26^.

As we were interested primarily in participants’ estimates of short- and long-term outcomes of different treatment alternatives, likelihood estimates at 0, 1 and 3 months were combined to create an average estimate for short-term outcomes. Combined estimates at 6, 9 and 12 months constituted long-term outcomes.

Estimates of surgical and non-surgical outcomes were analysed separately, as were the estimates of the likelihood of 1) engaging in normal activities (Activities), 2) experiencing pain (Pain), 3) experiencing depression (Depression), 4) experiencing complications (Complications). For each analysed surgical and non-surgical outcome, the full beta regression model included Condition (CABG, ColRec, Ortho), Timeframe (Short-term vs. Long-term), Group (Doctors vs. Patients) and all possible interactions as fixed factors, and participant ID as a random factor. Bonferroni correction was used to adjust for multiple comparisons, with *p*<.002 used as a significance cut-off point.

To investigate potential misalignment between doctors and patients in terms of the treatment choices made, participants decisions (Surgery vs. Non-surgical Alternative) were analysed using binomial generalized linear mixed models (GLMM), with Task (Estimation vs. Perspective), Group (Doctors vs. Patients) and Condition (CABG, ColRec, Ortho) and all possible interactions as fixed factors, and participant ID as a random factor. Nine participants had to be removed from this analysis due to data collection error, resulting in 100 separate choices being analysed.

The most parsimonious model was selected based on Akaike Information Criterion (AIC) from the set of models including all possible combinations of fixed factors. AIC values were calculated using the Maximum Likelihood (ML) method (check that). For post-hoc analyses Tukey HSD test was used as a method of adjusting *p* values for multiple comparisons using the lsmeans R package ^27^.

## Results

Results of the Likelihood Ratio Test of fixed effects for models of Activities, Pain, Depression, Complications and Choice are available in Supplementary Online Materials

### Likelihood of different outcomes when the surgical option is chosen: is there misalignment between patients and doctors?

Initial analysis of the surgical data revealed no significant effects of Condition (CABG, ColRec, Ortho), so for the final analysis the data was collapsed across conditions. Overall, patients were found to make more positive forecasts of surgical outcomes than doctors, particularly when the long-term outcomes were concerned, as demonstrated in Figure 1. They were found to forecast significantly higher likelihood of engaging in normal activities in the long-term (β=-1.09, SE=.20, *t*=-5.38, *p*<.001), lower likelihood of experiencing complications in the long-term (β=0.92, SE=.21, *t*=4.45, *p*<.001), and lower likelihood of experiencing depression in the in the short- and long-term (β=1.01, SE=.19, *t*=5.38, *p*<.001) after surgery. Estimates of the likelihood of experiencing pain were similar for patients and doctors.

**Figure 1.**
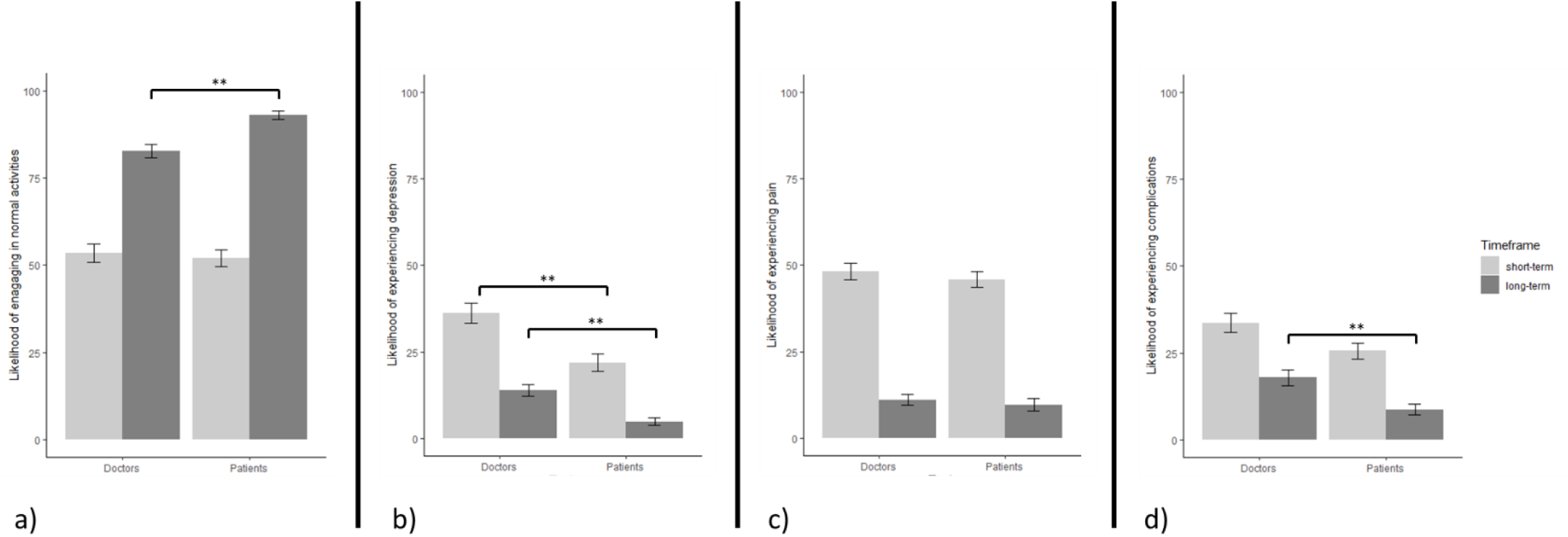
Likelihood estimates for different outcomes following surgery: a) engaging in normal activities, b) experiencing depression, c) experiencing pain, d) experiencing complications

### Likelihood of different outcomes when the non-surgical alternative is chosen: is there misalignment between patients and doctors?

Patients were found to make higher estimates of the likelihood of experiencing complications in the short-term when the non-surgical alternative was selected (β=-0.91, SE=.26, *t*=-3.50, *p*=.003) in comparison to doctors. No significant differences in estimates of the likelihood of engaging in normal activities or the likelihood of experiencing pain or depression were found, as indicated in Figure 2.

**Figure 2.**
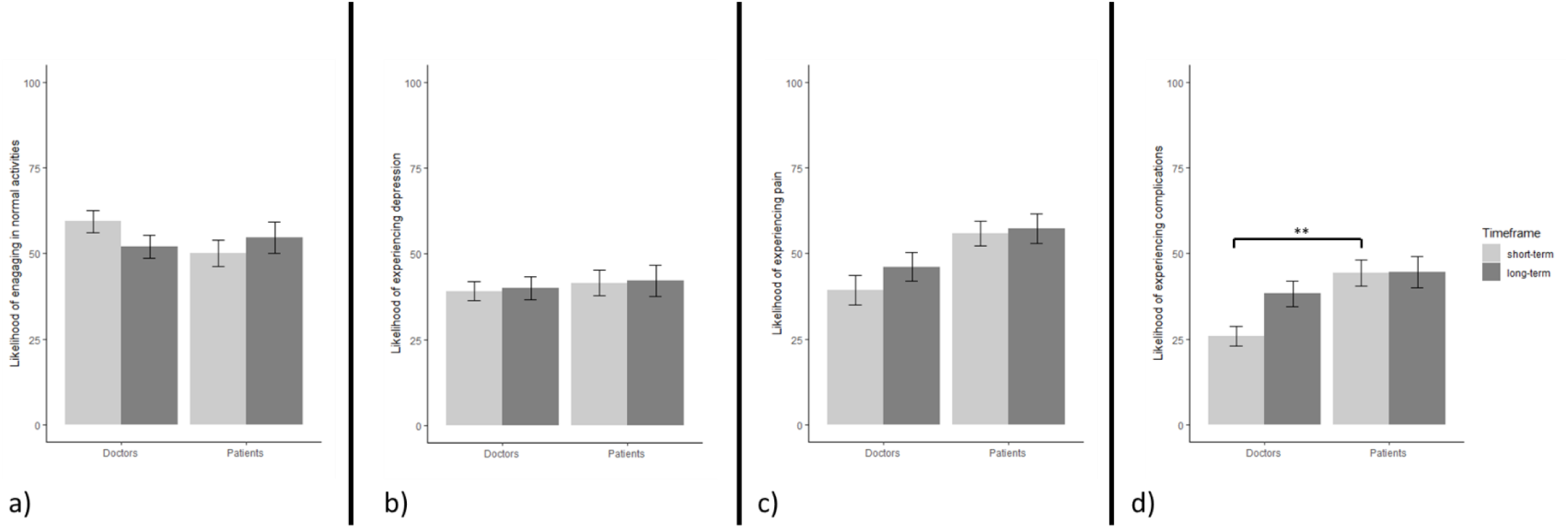
Likelihood estimates for different outcomes following the consultation in which surgery was declined: a) engaging in normal activities, b) experiencing depression, c) experiencing pain, d) experiencing complications

### Changing perspective: does it reduce the misalignment?

Overall, while an opportunity to adopt a perspective of the other side did lead to some adjustments in the estimates of treatment outcomes, it did not fundamentally reduce the misalignment between patients and doctors. Patients adopting doctors’ perspective were found to make lower estimates of the likelihood of experiencing complications in the short- and long-term following surgery (β=.86, SE=.19, *t*=4.50, *p*<.001), and lower estimates of the likelihood of engaging in normal activities in the short-term after surgery (β=0.62, SE=.18, *t*=3.46, *p*=.003) than actual doctors. No significant differences between the estimates of patients pretending to be doctors and actual doctors were found for pain and depression, as well as for non-surgical treatment outcomes.

In line with the trends observed in the Estimation task, doctors adopting patient’s perspective were found to be less positive about surgical outcomes than actual patients: estimating lower likelihood of engaging in normal activities in the long-term (β=-.89, SE=.21, *t*=-4.33, *p*<.001), and higher likelihood of experiencing depression (β=.54, SE=.19, *t*=2.86, *p*=.005) overall. For the non-surgical alternative, doctors adopting the patient perspective estimated lower likelihood of experiencing complications in the year following the consultation (β=-.61, SE=.24, *t*=-2.54, *p*=.01) than actual patients.

### Does the misalignment impact the choices that doctors and patients make?

Examination of the treatment choices participants made revealed that surgery was the most popular option in the ColRec (100% chose surgery) and Ortho (97% chose surgery) conditions, but less so in the CABG (64.3% chose surgery) condition. Since no participants chose the non-surgical alternative in the ColRec condition, a GLMM model with Condition as a fixed factor could not be estimated due to complete separation. For that reason, to explore if the differences in choices observed between conditions, Chi-Square Test of Independence (Estimation task) and Fisher’s Exact Test (Perspective task) were used with data collapsed across groups. Comparison of choices made in the Estimation task revealed that participants (both patients and doctors) were significantly more likely to choose alternative non-surgical treatment in the CABG condition than the Ortho (χ^2^(1,N=76)=11.79, *p*<.001) and ColRec condition (χ^2^(1,N=68)=11.41, *p*<.001). In the Perspective task, both patients and doctors were found to be significantly more likely to choose the non-surgical alternative in the CABG condition than the ColRec condition (*p*=.011, Fisher’s exact test), but not the Ortho condition.

To investigate the potential differences in choice behaviour between patients and doctors, and to explore changes in decisions from Estimation to Perspective task, binomial GLMM was used, with data collapsed across conditions. In the resulting analysis, patients were found to be significantly more likely to choose surgery when assuming the role of doctors than when making decisions as patients (β=-6.37, SE=1.92, *z*=-3.31, *p*=.005). Crucially, overall, no significant differences between the choices doctors and patients made were found. Majority of participants opted for surgery during both the Estimation task (88.7% of doctors and 80% of patients) and the Perspective task (82.2% of doctors, 90.1% of patients).

## Discussion

In this study, we attempted to identify the areas of potential misalignment in expectations between patients and doctors considering surgery, and the impact it has on treatment choice. The key finding was that while the expectations of patients and doctors differed considerably, the choices that they made were similar. Further, while an opportunity to adopt a perspective of the other side did lead to some adjustments in the estimates of treatment outcomes, it did not fundamentally reduce the misalignment between patients and doctors. In the following section we discuss the implications of this finding for the SDM approach, and the practical recommendations that follow from it.

Overall, our findings revealed a mismatch between the expectations of patients and doctors, which depended on the treatment option being considered. When forecasting surgical outcomes, patients were typically found to adopt a more positive outlook than doctors, particularly when considering long-term consequences. For non-surgical alternatives, where a mismatch was found between patients and doctors (i.e. in likelihood of experiencing complications), the estimates of patients were more negative than those of doctors. An intervention aimed at aligning the expectations of patients and doctors through perspective taking did little to reduce this mismatch, as patients remained significantly more positive about surgical outcomes and more negative about non-surgical outcomes than doctors, regardless of the perspective adopted. At the same time, while a misalignment between patients and doctors was detected, the treatment choices of the two groups did not differ. For choices, what mattered was the medical problem to contend with and the treatment options that were available.

Participants presented with a clinical scenario that described osteoarthritis of the hip or colorectal cancer had a strong preference for surgery. This was not observed to the same extent in the ischaemic heart disease scenario, where non-surgical alternative was selected 37% of the time. While the three scenarios differed in terms of overall prognosis and risks associated with surgery, these factors did not seem to be of primary concern when making decisions about treatment. For treatment choices, what mattered more was availability of an alternative treatment option that actively addressed the undesirable status quo. In the osteoarthritis and colorectal cancer scenarios, participants faced a choice between surgery and an alternative that could be construed as ‘doing nothing’ (management with medications, which has not been effective in the past; palliative care). In the ischaemic heart disease scenario an option of an active treatment (i.e. stents) was provided. This constituted ‘doing something’, so was a more attractive proposition to consider. Our results suggest that if forced to choose between ‘doing something’ (i.e. surgery) and ‘doing nothing’ (i.e. non-surgical treatment), it is likely that ‘doing something’ will be chosen, regardless of the personal characteristics of the decision-maker (e.g. whether they are a patient or a doctor, or what their expectations about outcomes are).

Studies on agency and control provide an explanation as to why such pattern of results was observed. In particular, findings show that in laboratory studies where participants are faced with conditions of uncertainty there is a strong preference to choose to act in order to reduce uncertainty, as well as enhance a sense of control over the situation^28, 29^. Under dynamic uncertainty, often experienced in medical settings, the outcomes (symptoms) change both as a result of actions taken (treatment), but also because of properties endogenous to the context (e.g. disease progression). In laboratory tasks that examine the way people learn in such circumstances, participants typically avoid a “do nothing” strategy in favour of making multiple interventions^30-32^. This occurs despite the fact that in learning environments which involve dynamic uncertainty of the kind described, taking a “do nothing” strategy early on is useful as a way of learning the dynamic properties. Taking up this strategy early helps to isolate one’s own causal impact from the changes that occur independently of those actions. Nonetheless, at the expense of this, commonly opting to act by continually intervening gives the impression of taking control. This phenomenon has also been observed in the real world contexts: where a desirable option is made the default (e.g. opt-out in organ donation register systems), when asked, people have a stronger preference for active choices systems (e.g. opt-in, mandated choice)^33^.

There is a strong theoretical precedent for asserting the psychological importance of active choice, because it is a means of engendering agentic experiences, and is ultimately adaptive^34^. Agentic experiences refer to situations where deliberative, purposeful actions are made, and when they are made, they enable a sense of agency and control^35^. Several theories spanning psychology outline mechanisms where active choice is also critical for wellbeing and self-esteem, as well as reducing experiences of uncertainty (Social Learning theory^35^, Reactance theory^36^, Self-determination theory^37^, Intention-action theory^38^, Rubicon model of action phases^39, 40^, Dynamic monitoring and control theory^28, 41^).

In terms of the practical implications of these findings for SDM in surgical context, our study highlights the importance of considering the decision-making problem that patients and doctors face. More specifically, we propose that there are three crucial components to the SDM process that need to be taken into account: there is a) the patient, b) the doctor, c) the decision to be made. Research on SDM focuses on the characteristics of the first two: their ability to convey and understand information, their knowledge, experience, goals and preferences^6, 42, 43^. So far little consideration has been given to the decision problem that patients and doctors face, and how it is framed. Decision-making literature demonstrates that how the choice alternatives are presented (e.g. which features are emphasised), can have an impact on the choice that is made^44, 45^. In the context of surgery, presenting this treatment as the only ‘active’ option that allows patients to maintain a sense of agency and control will make it likely that this option is chosen, despite the associated risks.

The propensity to opt for an ‘active’ treatment option to maintain the sense of agency and control has particularly important implications for high-risk patients considering surgery. As these patients are more likely to experience short-term and long-term complications following surgery^2, 3^, careful consideration of the potential consequences of this treatment and any alternatives is crucial to achieve a satisfactory decision. Such deliberation is unlikely to happen if the choice patients are facing is between ‘doing something’ (surgery) and ‘doing nothing’ (alternative treatment), as in such circumstances patients are likely to prioritise maintaining the sense of agency and control over exhaustive analysis of the pros and cons of different options. Based on the findings of this study, to encourage more in-depth processing of the information about treatment alternatives, we recommend that the non-surgical option is presented as “doing something”, a choice that leads to active disease management with a potential to bring about tangible benefits.

To our knowledge, this is the first study to directly compare outcome expectations and treatment decisions of high-risk individuals and doctors involved in surgery and perioperative care. Rigorous design, developed combining psychological and clinical expertise, allowed us to evaluate people’s perception of short- and long-term consequences of high-risk surgery, as well as the impact of these perceptions on actual choice of treatment, using online tools. Online design enabled consistent, efficient and safe data collection during the Covid-19 pandemic. It also gave us an opportunity to extend the geographical reach of our recruitment efforts and allowed us to protect participants from unnecessary stress by reaching people who, although suffering from several comorbidities, were not considering surgery at the time. Through our multidisciplinary approach, we were able to verify the usefulness of the perspective-taking in the SDM context and suggest other practical ways in which the objectives of the SDM can be achieved, without increasing the time or financial burden on the care providers.

As with any experiment exploring decision-making in hypothetical scenarios, a possibility remains that our findings would be somewhat different in real-life situations. Future studies should explore the impact of the framing of the choice alternatives (‘do something’ vs. ‘do nothing’) on the actual decisions made by patients and clinicians in the consultation rooms. Considering that our sampling focused on CCI scores, age (lay participants) and clinical role (doctors), future studies also need to establish if the same findings would be obtained from a larger, more representative sample. Finally, online delivery prevented us from establishing how effective the perspective-taking was in our task, i.e. how engaged our participants were when pretending to be patients/doctors. Further, in-person investigations would be needed to clarify if more immersive perspective-taking interventions would be more effective at aligning the expectations of patients and clinicians.

## Conclusions

Misalignment between patients and doctors in their expectations regarding the outcomes of surgical and non-surgical treatment constitutes a potential barrier to effective SDM in surgical context. Our study identified such misalignment, revealing that patients tend to be more positive about surgical outcomes, and to some extent more negative about non-surgical outcomes than doctors. This mismatch between patients and doctors remained even when an opportunity to adopt the perspective of the other side was provided. Despite that, both groups made very similar decisions regarding treatment. We propose that participants’ choices were guided primarily by the presence or absence of ‘active’, actionable non-surgical treatment alternatives that allow for a sense of agency and control to be maintained without resorting to surgery. In light of these findings, to improve SDM in surgical settings we advocate greater focus on the framing of the decision-making problem, particularly on how different treatment alternatives are presented to patients: as ‘active’ and capable of changing the status quo. Equalizing the surgical and non-surgical option in this way could potentially increase the willingness of both patients and doctors to consider the consequences of each option, allowing for an informed, patient-tailored decision to be made

## Data Availability

All data produced in the present study are available upon reasonable request to the authors

